# Safety and immunogenicity of a hybrid-type vaccine booster in BBIBP-CorV recipients: a randomized controlled phase 2 trial

**DOI:** 10.1101/2022.03.08.22272062

**Authors:** Nawal Al Kaabi, Yun Kai Yang, Li Fang Du, Ke Xu, Shuai Shao, Yu Liang, Yun Kang, Ji Guo Su, Jing Zhang, Tian Yang, Salah Hussein, Mohamed Saif ElDein, Sen Sen Yang, Wenwen Lei, Xue Jun Gao, Zhiwei Jiang, Xiangfeng Cong, Yao Tan, Hui Wang, Meng Li, Hanadi Mekki Mekki, Walid Zaher, Sally Mahmoud, Xue Zhang, Chang Qu, Dan Ying Liu, Jing Zhang, Mengjie Yang, Islam Eltantawy, Jun Wei Hou, Ze Hua Lei, Peng Xiao, Zhao Nian Wang, Jin Liang Yin, Xiao Yan Mao, Jin Zhang, Liang Qu, Yun Tao Zhang, Xiao Ming Yang, Guizhen Wu, Qi Ming Li

**Author notes:** These authors contributed equally: Nawal Al Kaabi, Yun Kai Yang, Li Fang Du, Ke Xu.

## Abstract

The emergence of severe acute respiratory syndrome coronavirus 2 (SARS-CoV-2) variants with immune escape ability raises the urgent need for developing cross-neutralizing vaccines against the virus. NVSI-06-08 is a potential broad-spectrum recombinant COVID-19 vaccine that integrates the antigens from multiple SARS-CoV-2 strains into a single immunogen. Here, we evaluated the safety and immunogenicity of NVSI-06-08 as a heterologous booster dose in adults previously vaccinated with the inactivated vaccine BBIBP-CorV in a randomized, double-blind, controlled, phase 2 trial conducted in the United Arab Emirates (NCT05069129). Three groups of healthy adults over 18 years of age (600 participants per group) who had administered two doses of BBIBP-CorV 4-6-month, 7-9-month and >9-month earlier, respectively, were vaccinated with either a homologous booster of BBIBP-CorV or a heterologous booster of NVSI-06-08. The primary outcome was immunogenicity and safety of booster vaccinations. The exploratory outcome was cross-reactive immunogenicity against multiple SARS-CoV-2 variants of concerns (VOCs). The incidence of adverse reactions was low in both booster vaccinations, and the overall safety profile of heterologous boost was quite similar to that of homologous boost. Heterologous NVSI-06-08 booster was immunogenically superior to homologous booster of BBIBP-CorV. Both Neutralizing and IgG antibodies elicited by NVSI-06-08 booster were significantly higher than by the booster of BBIBP-CorV against not only SARS-CoV-2 prototype strain but also multiple VOCs. Especially, the neutralizing activity induced by NVSI-06-08 booster against the immune-evasive Beta variant was no less than that against the prototype strain, and a considerable level of neutralizing antibodies against Omicron (GMT: 367.67; 95%CI, 295.50-457.47) was induced by heterologous booster, which was substantially higher than that boosted by BBIBP-CorV (GMT: 45.03; 95%CI, 36.37-55.74). Our findings showed that NVSI-06-08 was safe and immunogenic as a booster dose following two doses of BBIBP-CorV, which was immunogenically superior to homologous boost with another dose of BBIBP-CorV. Our study also indicated that the design of hybrid antigen may provide an effective strategy for broad-spectrum vaccine developments.

## Main

Through the great efforts of researchers worldwide, remarkable achievements have been made in developing effective vaccines against coronavirus disease-19 (COVID-19). As of January 31, 2022, a total of 10 vaccines have been authorized by World Health Organization (WHO) for emergency use^1^, and 4,084,470,843 individuals (52.1% of the population) in the world have been fully vaccinated^2^. These COVID-19 vaccines provide efficient protection against severe acute respiratory syndrome coronavirus 2 (SARS-CoV-2). However, the virus is continuously evolving, and a number of variants have emerged, some of which acquired immune escape capability.^3^ Due to the pandemic of SARS-COV-2 variants and the waning of immunity over time, breakthrough infections are growing rapidly^3,4^. The Technical Advisory Group on COVID-19 Vaccine Composition (TAG-CO-VAC) of WHO has recommended updating the composition of current COVID vaccines to develop multivalent or broad-protective vaccines against SARS-CoV-2 current and even future variants^5^.

Guided by structural and computational analysis of the receptor-binding domain (RBD) of SARS-CoV-2 spike protein, we have designed a mutation-integrated trimeric RBD (mutI-tri-RBD) as the antigen of a recombinant COVID-19 vaccine named NVSI-06-08 (Sinopharm)^6^. In mutI-tri-RBD, three heterologous RBDs, derived respectively from the prototype, Beta and Kappa SARS-CoV-2 strain, were connected end to end and co-assembled into a trimeric structure. By this way, mutI-tri-RBD serves as a hybrid antigen that integrates key mutations from multiple SARS-CoV-2 variants into a single protein. Pre-clinical studies have demonstrated that NVSI-06-08 elicited broader immune response against SARS-CoV-2 variants. The hybrid strategy has also been applied to HIV coronaviruses and influenza vaccine developments, and it has been proved that hybrid-type vaccine not only can improve immune response but also effectively expand the breadth of immunity^7-9^.

As of now, more than half of the world’s population has received at least one shot of COVID-19 vaccine^2^, and thus an effective broad-reactive vaccine is needed as a booster dose to strengthen and broaden immunity against SARS-CoV-2 variants. The inactivated vaccine BBIBP-CorV made by Sinopharm is one of the COVID-19 vaccines approved by WHO and has been used in many countries^10^. Given that large-scale populations worldwide have already administered two doses of BBIBP-CorV, in this trial, we evaluate the immunogenicity and safety of NVSI-06-08 as a heterologous booster dose, using homologous boost with BBIBP-CorV as control. As an exploratory study, the cross-reactive immunogenicity of the heterologous BBIBP-CorV/NVSI-06-08 prime-booster vaccination against SARS-CoV-2 variants of concerns (VOCs), including Omicron, was also assessed and compared with that of the homologous booster vaccination with BBIBP-CorV to illustrate the superior immunogenicity of NVSI-06-08 as a booster dose.

## Results

### participants

A total of 1833 healthy adults (≥18 years old) were enrolled, and these participants were classified into three groups with different prime-boosting intervals, i.e., 4-6 months, 7-9 months and >9 months. For each group, participants were randomly assigned to receive either a heterologous boost of NVSI-06-08 or a homologous boost of BBIBP-CorV. Among the enrolled participants, 1800 individuals completed booster vaccination, with 600 participants in each group (Fig. 1). The majority of the participants were Bangladeshis, Indian or Pakistanis, and more men than women participated in the trial. The demographic characteristics were broadly similar between heterologous and homologous boosting groups (Table 1 and Extended Data Table 1). The participants in two groups had similar age, sex, race, height and weight distributions. All the 1800 participants who had received the booster dose of vaccination were included in Safety Set (SS) for safety analysis. A total of 1678 participants who had no protocol deviations from the follow-up visits were included in Per-protocol set (PPS) for baseline analysis immunogenicity evaluation (Fig. 1).

**Fig. 1.**
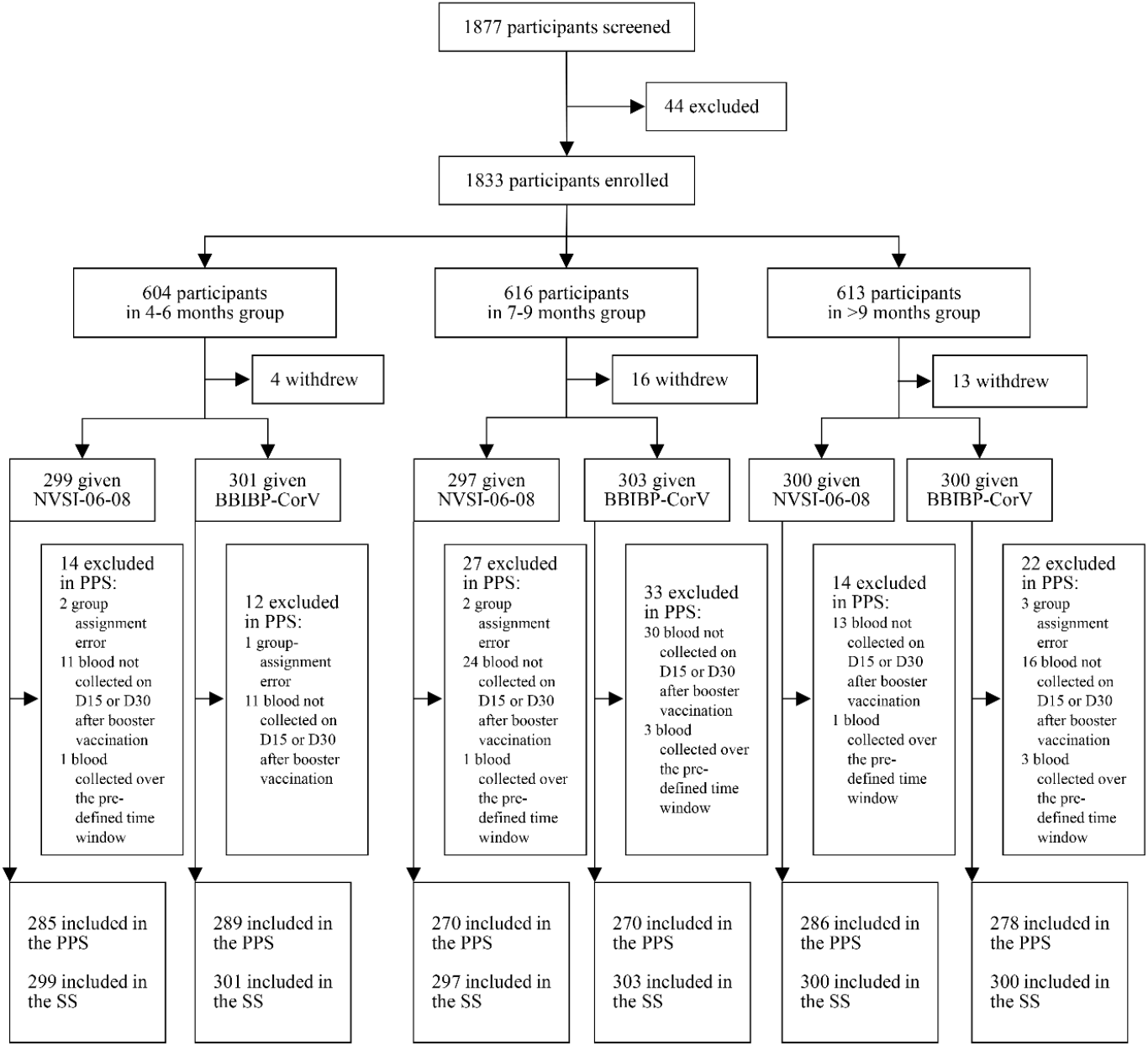
Randomization and analysis populations. A total of 1833 participants were enrolled, and 1800 received booster vaccinations. Participants were classified into three groups with different prime-boost intervals. The participants in each group were randomly assigned to receive a booster dose of eighter NVSI-06-08 or BBIBP-CorV. All the 1800 participants receiving booster vaccination were included in safety set (SS) for safety analysis. A total of 1678 participants who had no protocol deviations on follow-up visits were included in Per-protocol set (PPS) for immunogenicity analysis.

**Table 1.** Demographic characteristics of the participants.

|  | 4-6 months |  |  | 7-9 months |  |  | > 9 months |  |  |
| --- | --- | --- | --- | --- | --- | --- | --- | --- | --- |
|  | NVSI-06-08<br>(N=285) | BBIBP-CorV<br>(N=289) | P value | NVSI-06-08<br>(N=270) | BBIBP-CorV<br>(N=270) | P value | NVSI-06-08<br>(N=286) | BBIBP-CorV<br>(N=278) | P value |
| Age (years) |  |  |  |  |  |  |  |  |  |
| Mean (SD) | 35.47 (8.23) | 34.80 (8.15) | 0.326 | 34.79 (8.39) | 34.61 (8.25) | 0.802 | 37.16 (8.29) | 36.86 (7.85) | 0.657 |
| Median (IQR) | 35.13 (29.47-40.45) | 33.85 (29.60-39.84) |  | 33.84 (28.62-40.05) | 34.13 (28.46-40.15) |  | 36.41 (31.05-42.23) | 36.77 (31.72-41.46) |  |
| Min, Max | 19.65, 59.84 | 19.60, 64.84 |  | 19.16, 66.81 | 18.68, 60.49 |  | 18.96, 66.45 | 19.85, 60.85 |  |
| Age group, n(%) |  |  |  |  |  |  |  |  |  |
| 18-59 years | 285 (100.00) | 288 (99.65) | 0.320 | 269 (99.63) | 269 (99.63) | 1.000 | 284 (99.30) | 277 (99.64) | 0.579 |
| ≥60 years | 0 (0.00) | 1 (0.35) |  | 1 (0.37) | 1 (0.37) |  | 2 (0.70) | 1 (0.36) |  |
| Sex, n(%) |  |  |  |  |  |  |  |  |  |
| Male | 280 (98.25) | 286 (98.96) | 0.464 | 261 (96.67) | 257 (95.19) | 0.384 | 275 (96.15) | 274 (98.56) | 0.076 |
| Female | 5 (1.75) | 3 (1.04) |  | 9 (3.33) | 13 (4.81) |  | 11 (3.85) | 4 (1.44) |  |
| Height (cm) |  |  |  |  |  |  |  |  |  |
| Mean (SD) | 168.60 (7.59) | 168.69 (6.34) | 0.886 | 170.51 (7.30) | 169.57 (6.95) | 0.124 | 169.26 (7.73) | 169.24 (7.70) | 0.976 |
| Median (IQR) | 168.00 (164.00-173.00) | 169.00 (165.00-173.00) |  | 170.00 (166.00-175.00) | 170.00 (165.00-174.00) |  | 170.00 (165.00-174.00) | 170.00 (165.00-174.00) |  |
| Min, Max | 124.00, 198.00 | 147.00-189.00 |  | 149.00-187.50 | 144.40, 187.00 |  | 118.00, 189.00 | 106.00, 189.00 |  |
| Weight (kg) |  |  |  |  |  |  |  |  |  |
| Mean (SD) | 73.38 (13.18) | 71.86 (11.04) | 0.135 | 75.25 (13.35) | 74.73 (13.10) | 0.647 | 75.64 (13.36) | 73.33 (13.00) | 0.038 |
| Median (IQR) | 72.00 (64.00-82.00) | 71.30 (64.00-79.00) |  | 74.00 (66.00-83.00) | 73.00 (65.00-83.00) |  | 74.00 (67.00-84.00) | 72.00 (65.00-79.50) |  |
| Min, Max | 43.00, 121.00 | 42.00, 113.00 |  | 38.50, 136.00 | 48.00, 117.00 |  | 45.00, 156.00 | 45.00, 124.00 |  |
| BMI (kg/m<sup>2</sup>) |  |  |  |  |  |  |  |  |  |
| Mean (SD) | 25.82 (4.46) | 25.28 (3.77) | 0.114 | 25.85 (4.11) | 25.94 (3.98) | 0.789 | 26.39 (4.28) | 25.65 (4.75) | 0.052 |
| Median (IQR) | 25.51 (22.92-28.33) | 25.10 (22.49-27.74) |  | 25.53 (23.03-28.36) | 25.47 (22.85-28.58) |  | 25.75 (23.92-28.40) | 25.20 (22.94-27.86) |  |
| Min, Max | 16.59, 54.63 | 15.06, 35.16 |  | 15.62, 46.51 | 17.73, 39.03 |  | 17.63, 50.36 | 15.04, 69.42 |  |
| Pre-booster<br>neutralizing<br>antibody GMT<br>(95%CI) | 78.35 (67.10-91.48) | 67.28 (57.41-78.84) | 0.1771 | 51.98 (44.89-60.19) | 48.80 (42.03-56.66) | 0.5526 | 16.96 (14.56-19.74) | 17.47 (14.94-20.44) | 0.7870 |
| Pre-booster RBD-specific IgG GMC<br>(95%CI) | 113.71(96.45-134.05) | 95.53(81.53-111.94) | 0.1341 | 133.93(111.27-161.20) | 118.77(97.65-144.47) | 0.3810 | 120.45(96.96-149.63) | 116.05(91.23-147.62) | 0.8210 |

For the participants, baseline IgG concentrations and neutralizing antibody titers were quantified using a chemiluminescence enzyme immunoassay kit and the live-virus neutralization assay, respectively, before booster vaccination. The baseline antibody levels were statistically similar between the participants in heterologous and homologous boosting groups. Before boosting vaccination, the baseline IgG GMCs were similar in the participants among groups with different prime-boosting intervals. However, neutralizing GMTs in the participants from >9-month group were lower than from 7-9-month group, and those from 7-9-month group were also lower than from 4-6-month group, which demonstrated wanning of neutralizing immunity over time (Table 1). The decay of neutralizing immunity highlighted the need for booster vaccination to top-up the immune response.

### Safety

In the trial, 146 (16.29%) participants receiving NVSI-06-08 boost and 115 (12.72%) receiving BBIBP-CorV boost reported at least one solicited adverse reaction within 7 days after vaccination, and most of them were of grade 1 or 2. The overall incidence of solicited adverse reactions was low in both booster vaccinations (Fig. 2a,b and Extended Data Table 2). The occurrence of solicited local adverse reactions was quite similar between heterologous and homologous booster groups. All the reported local reactions were of grade 1 or 2, and most of them were injection-site pain (Fig. 2a and Extended Data Table 2). Solicited systemic adverse reactions reported by participants in heterologous boosting groups were also similar to those reported by homologous boosting groups. The reported systemic reactions were mostly of grade 1 or 2, and the most frequent reactions were headache, muscle pain, fatigue and fever. Grade 3 systemic reactions, including fever and muscle pain, were only observed in 0.45% participants of heterologous groups and 0.22% participants of homologous group, respectively. No grade 4 or above systemic reaction was found (Fig. 2b and Extended Data Table 2). The proportion of participants reporting unsolicited adverse reactions was also comparable between heterologous and homologous booster groups (5.36% vs 5.31%). The observed unsolicited reactions primarily included myalgia (0.56% vs 0.66%), fever (0.45% vs 0.55%) and cough (0.33% vs 0.88%), most of which were graded as level 1 or 2 (Extended Data Table 2). In both groups, no AESI and vaccination-related SAE was reported as of the time of this report. Overall, these data suggest that the heterologous boosting with a dose of NVSI-06-08 following two doses of BBIBP-CorV has a good safety profile, which were quite similar to homologous boosting with BBIBP-CorV.

**Fig. 2.**
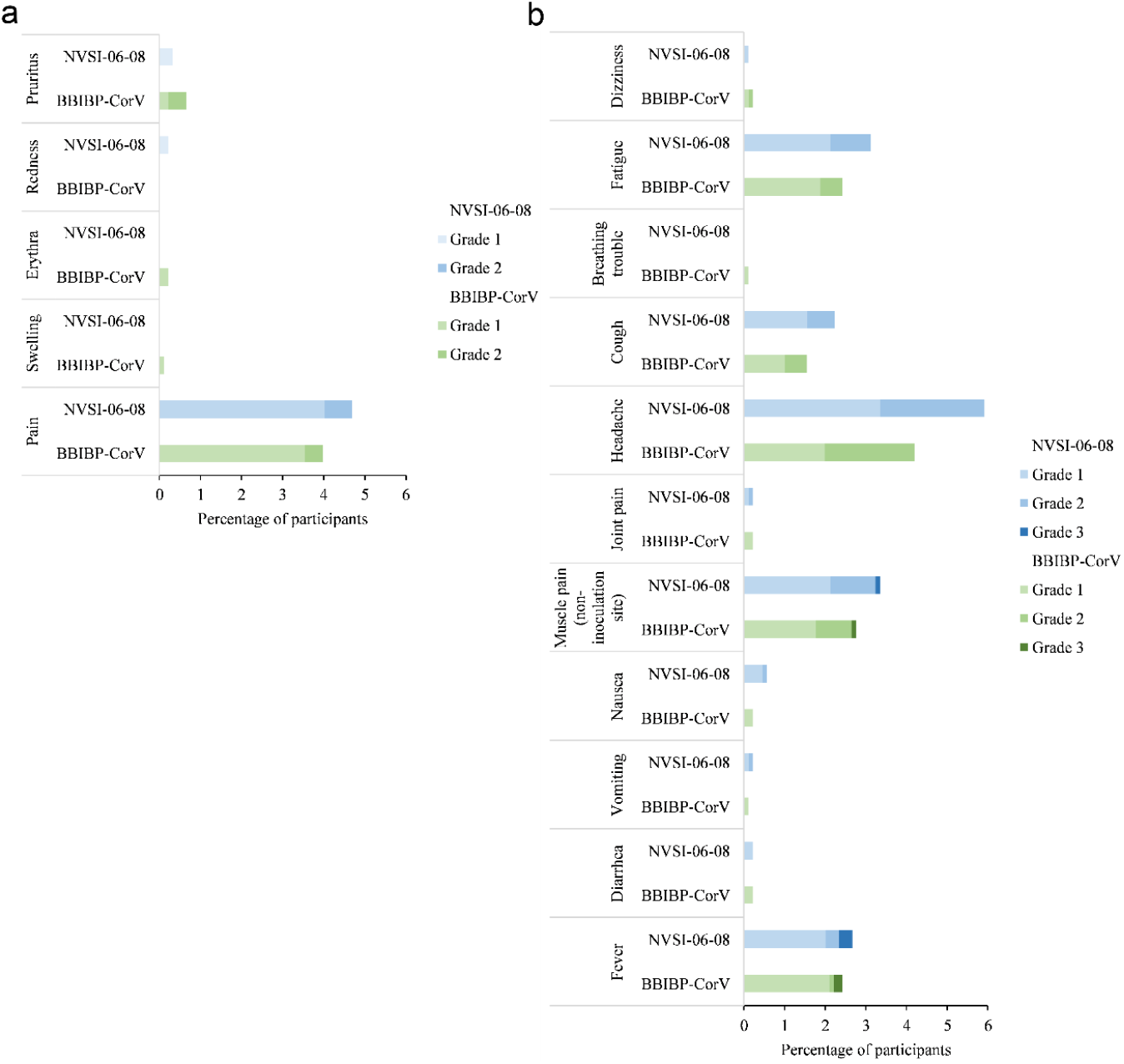
Incidence and severity of solicited adverse reactions after booster vaccinations with NVSI-06-08 and BBIBP-CorV, respectively. Incidence and severity of local (**a**) and systemic (**b**) adverse reactions after boosted with NVSI-06-08 were compared to those boosted with BBIBP-CorV. Adverse reactions are graded according to the relevant guidance of China National Medical Products Administration (NMPA).

### Immunogenicity against prototype virus

Significantly increase in neutralizing antibody titers against the prototype SARS-CoV-2 virus, detected by live-virus neutralization assays, were observed after both homologous and heterologous boosting vaccinations compared to the pre-boosting baseline values. However, the post-vaccination neutralizing antibody GMTs of heterologous boosting group were dramatically higher than those of homologous boosting groups. On day 15 post-vaccination, GMTs of neutralizing antibodies elicited by homologous BBIBP-CorV boost were increased by 2.93-fold (95%CI, 2.54-3.37) in 4-6-month group, 10.34-fold (8.78-12.19) in 7-9-month group and 21.44-fold (18.56-24.77) in >9-month group, respectively, compared to the pre-boosting baseline levels. Whereas, those elicited by heterologous NVSI-06-08 boost were improved by 40.10-fold (95%CI, 34.61-46.47), 94.42-fold (79.36-112.34) and 246.81-fold (207.02-294.26) in the three groups, respectively (Fig. 3a and Extended Data Table 3). Correspondingly, the 4-fold rise rates of neutralizing antibodies boosted by BBIBP-CorV were 22.84% (95%CI, 18.13%-28.12%), 75.19% (69.59%-80.22%), and 94.24% (90.82%-96.67%) in 4-6-month, 7-9-month and >9-month groups, while those boosted by NVSI-06-08 reached 93.68% (95%CI, 90.20%-96.21%), 98.15% (95.73%-99.40%) and 99.65% (98.07%-99.99%) in the three groups, respectively (Fig. 3b and Extended Data Table 3). Neutralizing antibody GMTs and 4-fold rise rates further increased on day 30 after booster vaccination, and the neutralizing responses induced by heterologous boost was also remarkably superior to those by homologous boost (*P*<0.0001). On day 30 post-vaccination, homologous boost of BBIBP-CorV led to 4.38-fold (95%CI, 3.81-5.05), 22.40-fold (19.19-26.15) and 46.26-fold (39.76-53.83) increases from baselines in neutralizing antibody GMTs for participants from 4-6-month, 7-9-month and >9-month groups, respectively, whereas much greater increases of 47.61-fold (95%CI, 41.17-55.06), 148.50-fold (126.60-174.19) and 441.11-fold (373.91-520.38) were obtained by heterologous boost of NVSI-06-08 (Fig. 3a and Extended Data Table 3). A similar increase trend was also observed in 4-fold rise rates. For 4-6-month, 7-9-month and >9-month groups, the 4-fold rise rates induced by homologous boost were 38.75% (95%CI, 33.11%-44.64%), 92.96% (95%CI, 89.23%-95.71%) and 98.20% (95.85%-99.41%), respectively, which were increased to 96.84% (95%CI, 94.09%-98.55%), 99.26% (97.35%-99.91%) and 100.00% (98.72%-100.00%) induced by heterologous boost (Fig. 3b and Extended Data Table 3). Our data indicate that heterologous boosting with NVSI-06-08 is dramatically more immunogenic than homologous boosting with BBIBP-CorV. Among three groups with different prime-boosting intervals, the post-vaccination neutralizing antibody levels in the participants of 7-9-month group were comparable to those of >9-month group, both of which were significantly higher than those of 4-6-month group. Especially, the neutralizing GMTs elicited by NVSI-06-08 in 7-9-month group reached as high as 7719.35 against wild-type SARS-CoV-2 virus. The results suggest that a booster dose with a prime-boosting interval over 6 months is immunogenically optimal.

**Fig. 3.**
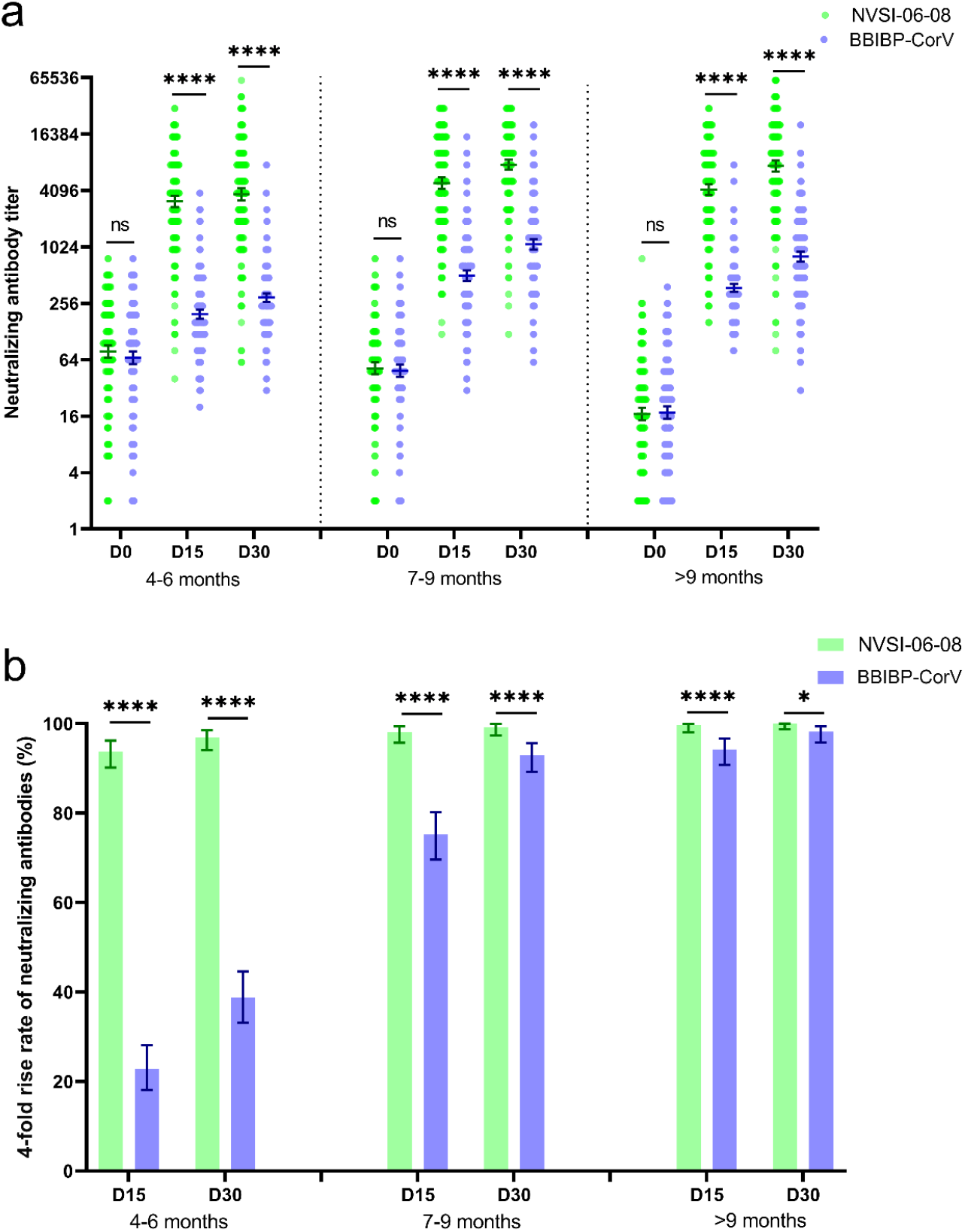
Neutralizing antibody levels against prototype SARS-CoV-2 before and 15 and 30 days after boosting. GMTs of neutralizing antibodies increased from baseline (day 0) to day 15 and 30 post-boosting elicited by heterologous NVSI-06-08 booster, compared with those induced by homologous BBIBP-CorV boosting (**a**). Correspondingly, the 4-fold rise rates of neutralizing antibodies on day 15 and 30 after boosting elicited by NVSI-06-08 booster, compared with those induced by BBIBP-CorV booster (**b**). Data are presented as GMTs and 95% CIs. *: *P*<0.05, ****: *P*<0.0001.

The immunogenic superiority of heterologous NVSI-06-08 booster to homologous BBIBP-CorV booster was also confirmed by anti-RBD IgG response. In line with the neutralizing antibody titers, both homologous and heterologous boosting vaccinations significantly improved the RBD-specific IgG antibody levels. However, heterologous booster induced dramatically higher IgG GMCs than homologous booster in all groups with different boosting intervals. Compared to pre-boosting baselines, anti-RBD IgG GMCs were increased by 2.56-3.58 folds at 15 days after vaccination in the groups boosted with BBIBP-CorV, whereas, much higher 49.15 to 62.62-fold increases were observed in the groups boosted with heterologous NVSI-06-08 (Fig. 4a and Extended Data Table 4). Similarly, 4-fold rise rates elicited by heterologous booster were significantly higher than those by homologous booster in all groups with different boosting intervals (90.56%-96.14% vs 20.76%-31.65%, *P*<0.0001) (Fig. 4b and Extended Data Table 4). At 30 days after boosting vaccination, similar results were obtained. IgG antibody GMCs boosted by NVSI-06-08 increased from baseline by 44.24-fold (95%CI, 37.82-51.75) in 4-6-month group, 36.94-fold (30.65-44.52) in 7-9-month group and 41.18-fold (33.25-51.01) in >9-month group, respectively, which were remarkably greater than 2.24-fold (95%CI, 1.94-2.58), 2.45-fold (2.04-2.93) and 2.31-fold (1.89-2.82) boosted by BBIBP-CorV (*P*<0.0001) (Fig. 4a and Extended Data Table 4). Correspondingly, the 4-fold rise rates induced by heterologous booster were 95.44% (95%CI,92.33%-97.55%), 92.22% (88.36%-95.12%) and 88.11% (83.79%-91.62%), which were much higher than 17.30% (13.12%-22.16%), 22.96% (18.08%-28.45%) and 21.94% (17.22%-27.27%) induced by homologous booster (Fig. 4b and Extended Data Table 4).

**Fig. 4.**
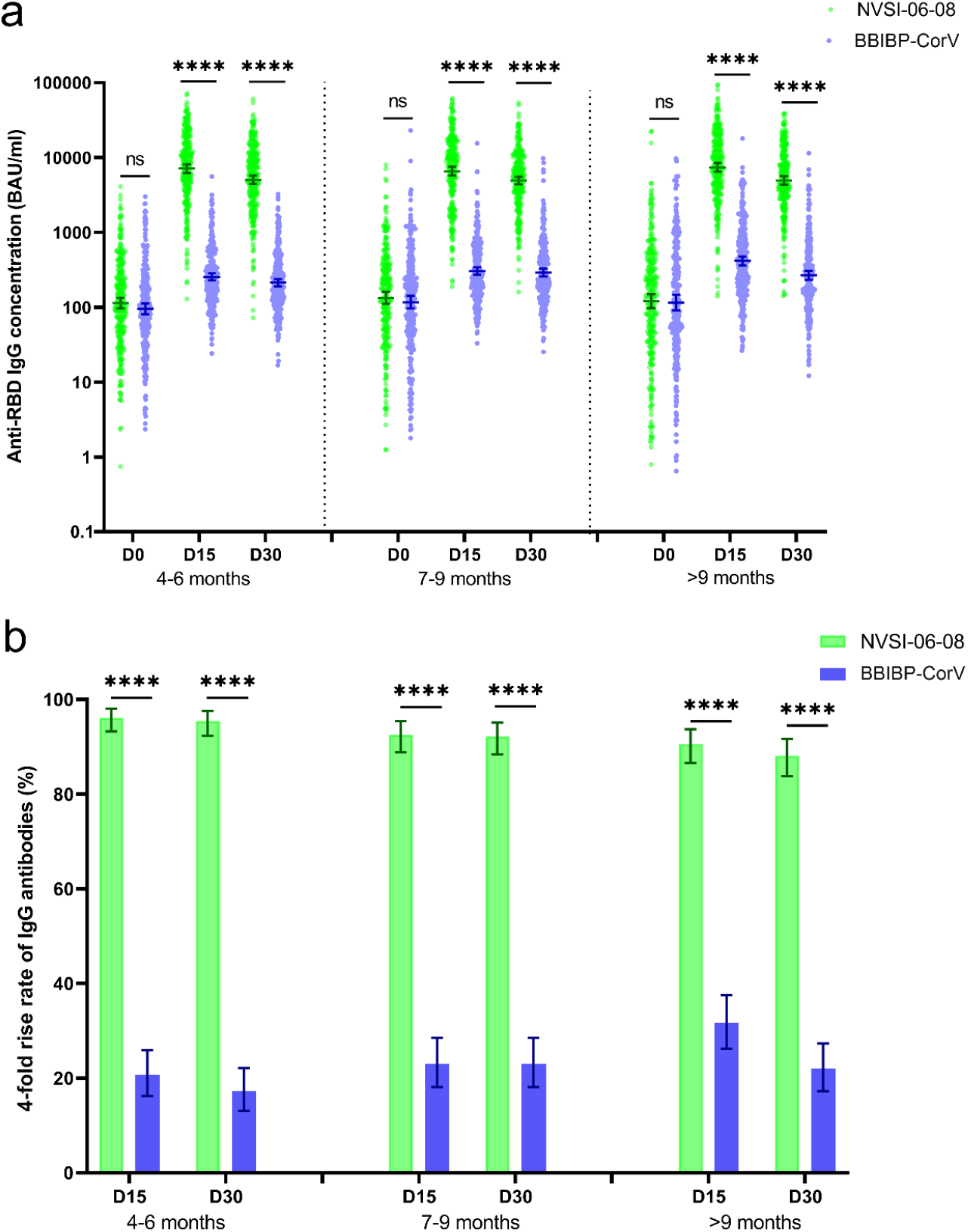
RBD-binding IgG antibody levels against prototype SARS-CoV-2 before and 15 and 30 days after boosting. GMCs of RBD-binding IgG antibodies increased from baseline (day 0) to day 15 and 30 post-boosting elicited by heterologous NVSI-06-08 booster, compared with those induced by homologous BBIBP-CorV boosting (**a**). Correspondingly, the 4-fold rise rates of IgG antibodies on day 15 and 30 after boosting elicited by NVSI-06-08 booster, compared with those induced by BBIBP-CorV booster (**b**). Data are presented as GMCs and 95% CIs. *: *P*<0.05, ****: *P*<0.0001.

### Neutralizing antibody response against Omicron and other VOCs

NVSI-06-08 was designed as a hybrid-type vaccine with broader neutralizing profiles, the cross-reactive immunogenicity of heterologous NVSI-06-08 booster against multiple SARS-CoV-2 VOCs, including Omicron, was evaluated as an exploratory study. A total of 200 serum samples, collected on day 15 post-boosting, from the participants with sequential enrollment numbers in 7-9-month group (99 participants receiving heterologous boost and 101 receiving homologous boost) were used in the cross-neutralizing activity tests.

In participants receiving the homologous boost of BBIBP-CorV, compared to the neutralizing antibody level against prototype SARS-CoV-2 strain [GMT: 460.39 (95%CI, 369.08-574.29)], the anti-Omicron neutralizing GMT significantly reduced to 45.03 (95%CI, 36.37-55.74), indicating distinct escape of Omicron variant from the immunity given by BBIBP-CorV. Our results are consistent with other studies^11-14^. By comparison, in participants receiving the heterologous boost of NVSI-06-08, the neutralizing antibody GMT against Omicron still maintained at a high level of 367.67 (95%CI, 295.50-457.47), which was much higher than that induced by homologous boost (Fig. 5 and Extended Data Table 5). Given that Omicron-specific vaccine is not yet available, a booster dose of NVSI-06-08 may serve as a possible choice against Omicron for BBIBP-CorV recipients.

**Fig. 5.**
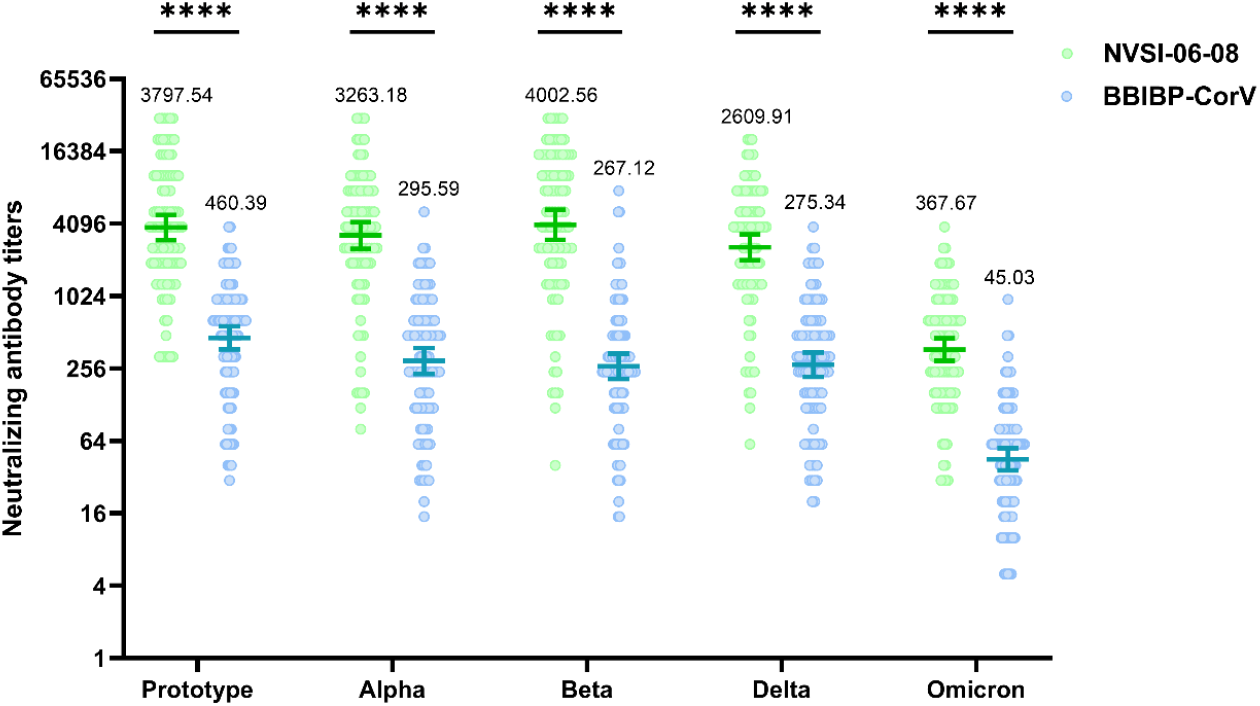
Cross-neutralizing antibody titers against SARS-CoV-2 prototype stain and several VOCs, including Alpha, Beta, Delta and Omicron, elicited by heterologous NVSI-06-08 booster, compared with those elicited by homologous BBIBP-CorV booster. A subset of 200 serum samples, collected on day 15 post-boosting, from the participants with sequential enrollment numbers in 7-9-month group (99 participants receiving a booster dose of NVSI-06-08 and 101 receiving a third dose of BBIBP-CorV) were tested using live-virus neutralization assay. Data are presented as GMTs and 95% CIs. The GMT values are given on the graph. ****: *P*<0.0001.

We also evaluated the neutralizing antibody response against other several SARS-CoV-2 VOCs, including Alpha, Beta and Delta. All the tested VOCs were significantly less sensitive to the neutralization induced by BBIBP-CorV booster. However, the neutralizing antibody response against Alpha and Beta variants offered by NVSI-06-08 booster was comparable to that against prototype strain. For all the tested variants, heterologous booster induced substantially greater neutralizing antibody levels than homologous booster. The GMTs of neutralizing antibodies boosted by NVSI-06-08 were 11.04-fold, 14.98-fold and 9.48-fold higher than those boosted by BBIBP-CorV against Alpha, Beta and Delta variants, respectively (Fig. 5 and Extended Data Table 5). NVSI-06-08 boosting vaccination is immunogenically superior to homologous BBIBP-CorV boosting against not only prototype SARS-CoV-2 strain but also other immune-evasive VOCs, including Omicron.

## Discussion

Findings from this trial show that the heterologous prime-boosting vaccination with one dose of NVSI-06-08 following two doses of BBIBP-CorV was safe, tolerant and immunogenic in healthy adults. The heterologous prime-boosting regimen with BBIBP-CorV/NVSI-06-08 was immunogenically superior to homologous BBIBP-CorV boost. The neutralizing antibody GMTs elicited by heterologous boost were 9.72-15.96-fold higher than those elicited by homologous boost (GMT, 3141.92-4908.34 versus 196.89-504.76) on day 15 post-boosting, and 7.06-12.65-fold higher (GMT, 3730.18-7719.35 versus 294.96-1093.26) on day 30 post-boosting. Compared to the peak value of neutralizing antibody level (GMT, 282.7) after priming vaccination with two doses of BBIBP-CorV as reported in the previous study^15^, the neutralizing GMT was improved 1.04-3.87-fold on day 30 after a booster dose of BBIBP-CorV, and more remarkably 13.19-27.31-fold after a heterologous booster dose of NVSI-06-08. Multiple lines of evidence have demonstrated the neutralizing antibody titers are highly correlated with protective efficacy against the infection of SARS-CoV-2^16-18^. The improvement of neutralizing antibody GMT is an indicator for enhancement of protective efficacy offered by the vaccine. Our findings indicate that a heterologous boost with NVSI-06-08 following prime vaccination with BBIBP-CorV could provide stronger protection against SARS-CoV-2 than a third dose of BBIBP-CorV.

The incidence of adverse reactions was low in both heterologous and homologous boosting vaccinations, and most of the reported local and systemic adverse reactions were of grade 1 or 2. The overall safety profile of heterologous boost was quite similar to that of homologous boost, which was also comparable to the safety of the priming with two doses of BBIBP-CorV as reported previously^15^. A booster dose did not distinctly increase the risk of more serious side effects. The antigen of NVSI-06-08 was designed to realize trimerization of RBDs without introducing any exogenous sequence, which facilitated the cross-links of B cell receptors but without introducing additional safety risks. In the vaccine, aluminum adjuvant was used, whose safety has been verified for a long time. All these features contributed to the high safety profile of NVSI-06-08.

NVSI-06-08 was designed as a hybrid-type COVID-19 vaccine with broad protection potential, which integrated multiple antigens into a single molecule without introduction any exogenous linker. Our studies show that booster vaccination of NVSI-06-08 elicited potent cross-neutralizing response against various SARS-CoV-2 VOCs. Especially, the neutralizing activity induced by NVSI-06-08 booster against the highly immune-evasive Beta variant was no less than that against the prototype strain, which demonstrated the immunological superiority of hybrid-type vaccine. Although the immunity offered by NVSI-06-08 booster was less effective against Omicron variant as compared to that against prototype virus, the anti-Omicron neutralizing GMT still maintained at a considerable level of 367.67 (95%CI, 295.50-457.47) on day 15 after booster vaccination, which was substantially higher than that boosted by BBIBP-CorV. According to the increasing trends in immune response, a greater anti-Omicron neutralizing GMT can be obtained on day 30 post-boost. Given that Omicron-specific vaccine is not yet available, the prime-boost vaccination with inactivated COVID-19 vaccine plus NVSI-06-08 may be an optional strategy against the pandemic of Omicron. The RBD of Omicron harbors numerous mutations, most of which were not integrated into the antigen of NVSI-06-08. We think that update of the vaccine to incorporate the Omicron-carrying mutations into the immunogen should induce better immunogenicity against Omicron variant. This strategy has been recently validated in animal experiments by our group (data not shown). Previous studies have indicated that hybrid-type immunogen could elicit superior B cell responses both in quantity and quality in comparison with the homologous immunogens. The superior immunogenicity of heterologous immunogen may be attributed to the avidity advantage to cross-reactive B cells^9^. Our studies provide a promising method for broad-spectrum COVID-19 vaccine development.

Considering that inactivated COVID-19 vaccines have been inoculated in large-scale populations worldwide, and the immunity offered by the vaccine decays obviously over time^19^, it is of significance to choose a preferred vaccine as a booster dose to restore and even top-up the immunity against SARS-CoV-2. Our studies provided a quite effective booster strategy for the inactivated vaccine recipients. The high level of neutralizing antibodies induced by heterologous BBIBP-CorV/NVSI-06-08 vaccination could alleviate the waning of immunity and facilitate the formation of immune barrier, which then may suppress virus mutations.

Our findings show that the prime-boost vaccination with an interval over 6 months was immunogenically superior to the interval of 4-6 months. Many studies have revealed that prime-boost interval has significant influence on the immune efficacy of COVID vaccines^20,21^. Our results are consistent with those previously reported studies. An over-6-month interval may facilitate better maturation of memory cells, and improve binding affinity and production level of antibodies.

This trial has several limitations. Firstly, among participants, the proportion of elderly persons aged ≥60 years was low, and the immune response of the BBIBP-CorV/NVSI-06-08 booster strategy in older population should be further assessed in the future. Secondary, the number of male participants were much larger than female, and the data may not well reflect the effect in women.

In summary, heterologous prime-boosting vaccination with BBIBP-CorV plus NVSI-06-08 was well tolerated and immunogenic against not only SARS-CoV-2 prototype strain but also the VOCs including Omicron. It was immunogenically superior to the booster with a third dose of BBIBP-CorV. Our study provides a preferred strategy to top up the immunity in BBIBP recipients against SARS-CoV-2 and its variants. The findings also implied that hybrid-type vaccine could induce potent and broad immune activities, which may provide an effective strategy for broad-spectrum vaccine developments.

## Data Availability

The clinical trial is still ongoing, and the data will be available when the trial is complete upon request to the corresponding author Qi Ming Li. After proposals are approved, data can be shared through secure online platforms.

## Methods

### Trial design and participants

We conducted a randomized, double-blind, controlled phase 2 trial to evaluate the immunogenicity and safety of NVSI-06-08 (Sinopharm) as a booster dose following a primary series of BBIBP-CorV (Sinopharm). Trial participants included three groups of healthy adults aged ≥18 years who had received two doses of BBIBP-CorV 4-6 months, 7-9 months and >9 months before, respectively. Participants were enrolled after undergoing a health screening by inquiry, medical history review and physical examination. Confirmed, suspected or asymptomatic COVID-19 cases, individuals with a history of SARS or MERS infections, and those vaccinated with any other COVID-19 vaccines were excluded. The detailed inclusion and exclusion criteria are available at ClinicalTrials.gov (NCT05069129).

### Trial oversight

The trial protocol was reviewed and approved by Abu Dhabi Health Research and Technology Ethics Committee. China National Biotec Group Co., Ltd. (CNBG) of Sinopharm was the regulatory sponsor of the trial. The trial was funded by Lanzhou Institute of Biological Products Co., Ltd (LIBP) of Sinopharm and Beijing Institute of Biological Products Co., Ltd (BIBP) of Sinopharm. National Vaccine and Serum Institute (NVSI) of Sinopharm and China National Biotec Group Co., Ltd. (CNBG) of Sinopharm designed the trial, performed the analyses and interpreted the data. All authors had full access to study data and the corresponding authors were responsible for the decision to submit the manuscript.

### Studied vaccines

NVSI-06-08 is a potential broad-spectrum recombinant COVID-19 vaccine, using a hybrid mutI-tri-RBD as the antigen, which was developed by the National Vaccine and Serum Institute (NVSI) of Sinopharm and manufactured by Beijing Institute of Biological Products Co., Ltd (BIBP) of Sinopharm and Lanzhou Institute of Biological Products Co., Ltd (LIBP) of Sinopharm in accordance with good manufacturing practice (GMP). One dose of NVSI-06-08 contains 20 μg antigen and 0·3 mg aluminum hydroxide adjuvant. BBIBP-CorV is an inactivated vaccine produced by Beijing Institute of Biological Products Co., Ltd (BIBP) of Sinopharm, which has been approved by WHO for emergency use and applied in many countries worldwide.

### Procedures

In each of the three groups with different prime-boost intervals, the participants were randomly assigned to receive either a heterologous boost of NVSI-06-08 or a homologous boost of BBIBP-CorV. After booster vaccination, participants were observed at study site for 30 min to identify immediate adverse reactions. Solicited adverse events (AEs) were recorded for 7 days and unsolicited AEs for 30 days post-vaccination. Serious adverse events (SAEs) and adverse events of special interest (AESIs) were collected up to 6 months after full course of immunization. The grades of local and systemic adverse events were determined according to the relevant guidance of China National Medical Products Administration (NMPA).

### Study outcomes

The primary immunogenicity outcome was the neutralizing response on 15 days and 30 days after booster vaccination, by evaluation of the geometric mean titers (GMTs) of neutralizing antibodies and the corresponding 4-fold rise rate (i.e., post-/pre-boost ≥4). The secondary immunogenicity outcome was geometric mean concentrations (GMCs) of IgG antibodies and the corresponding 4-fold rise rate. The safety outcome was occurrence and severity of any adverse reactions within 30 days post-boost. As an exploratory study, the immunogenicity of booster vaccination against SARS-CoV-2 variants of concerns (VOCs), including Omicron, was also evaluated in a subset of participants from 7-9-month group.

### Laboratory analyses

Spike receptor-binding domain (RBD)-specific IgG antibodies against the prototype SARS-CoV-2 strain was measured using a commercially available magnetic particle-based chemiluminescence enzyme immunoassay kit purchased from Bioscience (Chongqing) Biotechnology Co. (approved by the China National Medical Products Administration; approval numbers 20203400183). The IgG antibody detections were carried out on an automated chemiluminescence detector (Axceed 260) according to the manufacturer’s instructions. The reference calibrator used in the kit can be traced back to WHO International Standard First WHO International Standard for anti-SARS-CoV-2 immunoglobulin (human) NIBSC code:20/136.

Neutralizing antibody titers against prototype strain and VOCs, including Alpha, Beta, Delta and Omicron, were evaluated using live-virus neutralization assay as described in previous studies.^6^ In brief, serum samples were heat-inactivated at 56 °C for 30 min, and then serially diluted by 2 fold starting from 1:4 (in detection of neutralizing antibodies against prototype SARS-CoV-2 strain) or 1:10 (in detection of neutralizing antibodies against VOCs) dilution. The diluted serum was mixed with an equal volume of 100 TCID_50_ of SARS-CoV-2 live virus and incubated at 37 °C for 2 h. After that, the Vero cell suspension with a density of 1.5-2×10^5^ cells per mL was added into the serum-virus mixture, and then the plates were incubated at 37 °C for 3 to 5 days. Both cell-only and virus-only wells were also set as controls. Neutralizing antibody titer was determined as the reciprocal of the serum dilution for 50% protection against viral infection to the cell. The titer for the serum below the limit of detection was set to half value of the detection limit. The live-virus neutralization assays were performed in the BSL3 facility of National Institute for Viral Disease Control and Prevention, Chinese Center for Disease Control and Prevention (China CDC), Beijing, China. SARS-CoV-2 live viruses of the prototype (QD-01), Alpha (BJ-210122-14), Beta (GD84), Delta (GD96) and Omicron (NPRC2.192100003) strains were used in the assays. Both RBD-binding IgG detection and live-virus neutralization assays were carried out in a blinded manner.

### Statistical analysis

The sample size of participants was determined by the expected difference between groups, predefined noninferiority margin, intended power, significance level and estimated dropout rate. Assuming that the 4-fold rise rate after booster vaccination achieve 85%, 208 participants in each arm will be required to have 80% power to conclude non-inferiority with margin of -10% and one-sided significance level of 2.5% using Miettinen and Nurminen method. If equal GMT after booster immunization is assumed, and standard deviation of GMT after log10 transformation is considered to be 0.7, 250 subjects in each arm will be required to have 80% power to conclude non-inferiority with margin of 2/3 and one-sided significance level of 2.5%. Then, considering about 15%∼20% drop-out rate, 600 participants are required in each of the three groups (4-6-month, 7-9-month and >9-month groups), and 1800 subjects in total are planned to enroll.

Baseline characteristics were analyzed based on the participants who had no protocol deviations. Student’s t-test and Chi-square test were used, respectively, for the comparison of continuous and categorical characteristics between groups. Safety was analyzed based on the safety set (SS) that included the participants receiving booster vaccination. Safety analysis results were presented as counts and percentages of adverse reactions, along with the associated 95% confidence intervals (CIs) calculated by Clopper-Pearson method. The differences in safety between heterologous and homologous boosting groups were analyzed using Fisher’s exact test. Immunogenicity was evaluated based on the Per-protocol set (PPS), which included the participants without protocol deviation, and both the GMCs of RBD-specific IgG and GMTs of neutralizing antibodies were computed along with the associated Clopper-Pearson 95% CIs. According to the pre-booster and post-booster values, the fold rises in IgG GMCs and neutralizing antibody GMTs, as well as the associated 95% CIs, were calculated. In addition, the 4-fold rise rates of IgG and neutralizing antibodies along with 95%CIs were also computed. RBD-specific IgG concentrations and neutralizing antibody titers between heterologous and homologous boosting groups were compared using grouped t-test after log transformation. The fold rises in IgG GMC and neutralizing GMT between two groups were also compared using grouped t-test. 4-fold rise rates between heterologous and homologous booster groups were compared by Fisher’s exact test.

## Code availability

All codes that produced the results are available upon request to the corresponding author Qi Ming Li with a scientifically sound proposal.

## Acknowledgements

We thank Prof. Guoyong Yuan from the University of Hong Kong for providing SARS-CoV-2 Omicron virus used in cross-reactive neutralizing antibody detection assays.

## Author contributions

N.A.K. was the chief investigator. N.A.K., Q.M.L., Y.T.Z., X.M.Y., Y.K.Y., JingZ.(NVSI) and Y.K. designed the trial and study protocol. Q.M.L., JingZ.(NVSI), Y.L. and J.G.S. designed the recombinant vaccine NVSI-06-08. Q.M.L., Y.T.Z. and X.M.Y. were responsible for the organization and supervision of the project. Q.M.L., JingZ.(NVSI), Y.L., X.J.G. and X.Y.M. provided NVSI-06-08 vaccine. H.W. and JinZ. provided the inactivated vaccine BBIBP-CorV. X.M.Y., Y.T.Z., I.E. and Y.K.Y. contributed to project management. T.Y., Y.K., M.L., L.Q., W.Z., P.X., X.Z., C.Q., D.Y.L. and S.S.Y. participated in the implementation of the trial. H.M.M., Z.N.W. and J.L.Y. conducted sample collection and processing. G.W., K.X., W.L., JingZ.(China CDC), M.Y. and S.M. carried out the serum tests. G.W., K.X., Y.L. L.F.D., J.W.H. and Z.H.L. contributed to the development of serum testing method and manuscript preparation. S.H. and M.S.E. performed the clinical data gathering, analysis and operation. Z.J., X.C. and Y.T. performed statistical analysis of the data. Q.M.L., JingZ.(NVSI), Y.L., G.W., Y.K.Y. and K.X. performed data analysis and interpretation. J.G.S., JingZ.(NVSI), L.F.D., Y.L., K.X., S.S. and S.S.Y. contributed to the writing of the manuscript. Q.M.L. and G.W. oversaw final manuscript preparation. All authors approved the final manuscript.

## Additional information

**Extended data**

**Supplementary information**

**Extended Data Table 1.**
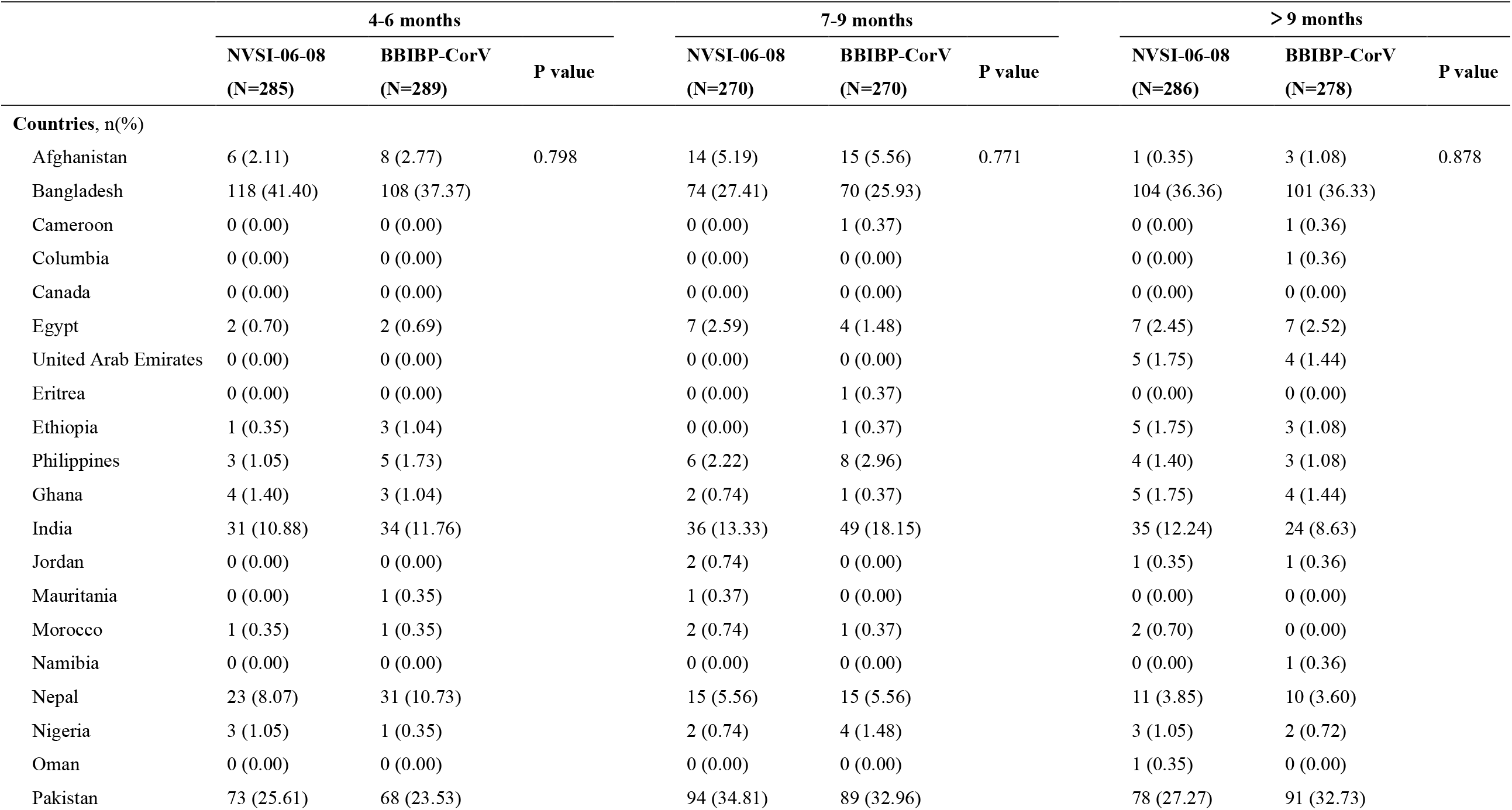
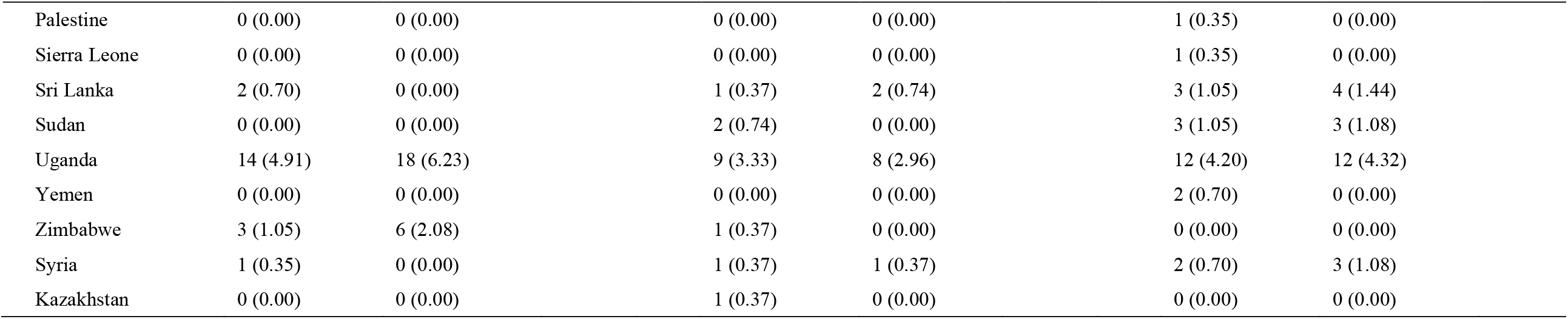
Baseline characteristic for the nationality of the participants.

**Extended Data Table 2.** Adverse reactions within 7 days of booster vaccination.

|  | NVSI-06-08<br>(N=896) | BBIBP-CorV<br>(N=904) | Total<br>(N=1800) | P value* |
| --- | --- | --- | --- | --- |
| **Solicited adverse reactions within 0-7 days** | 146 (16.29) | 115 (12.72) | 261 (14.50) |  |
| Grade 1 | 121 (13.50) | 94 (10.40) | 215 (11.94) |  |
| Grade 2 | 44 (4.91) | 35 (3.87) | 79 (4.39) |  |
| Grade 3 | 4 (0.45) | 2 (0.22) | 6 (0.33) |  |
| **Injection site adverse reactions** | 46 (5.13) | 42 (4.65) | 88 (4.89) |  |
| Grade 1 | 40 (4.46) | 37 (4.09) | 77 (4.28) |  |
| Grade 2 | 6 (0.67) | 8 (0.88) | 14 (0.78) |  |
| Pain | 42 (4.69) | 36 (3.98) | 78 (4.33) | 0.4888 |
| Grade 1 | 36 (4.02) | 32 (3.54) | 68 (3.78) | 0.6225 |
| Grade 2 | 6 (0.67) | 4 (0.44) | 10 (0.56) | 0.5460 |
| Swelling | 0 | 1 (0.11) | 1 (0.06) | 1.0000 |
| Grade 1 | 0 | 1 (0.11) | 1 (0.06) | 1.0000 |
| Erythra | 0 | 2 (0.22) | 2 (0.11) | 0.4997 |
| Grade 1 | 0 | 2 (0.22) | 2 (0.11) | 0.4997 |
| Redness | 2 (0.22) | 0 | 2 (0.11) | 0.2476 |
| Grade 1 | 2 (0.22) | 0 | 2 (0.11) | 0.2476 |
| Pruritus | 3 (0.33) | 6 (0.66) | 9 (0.50) | 0.5068 |
| Grade 1 | 3 (0.33) | 2 (0.22) | 5 (0.28) | 0.6858 |
| Grade 2 | 0 | 4 (0.44) | 4 (0.22) | 0.1246 |
| **Systemic adverse reactions** | 111 (12.39) | 84 (9.29) | 195 (10.83) |  |
| Grade 1 | 89 (9.93) | 65 (7.19) | 154 (8.56) |  |
| Grade 2 | 39 (4.35) | 30 (3.32) | 69 (3.83) |  |
| Grade 3 | 4 (0.45) | 2 (0.22) | 6 (0.33) |  |
| Fever | 24 (2.68) | 22 (2.43) | 46 (2.56) | 0.7671 |
| Grade 1 | 18 (2.01) | 19 (2.10) | 37 (2.06) | 1.0000 |
| Grade 2 | 3 (0.33) | 1 (0.11) | 4 (0.22) | 0.3724 |
| Grade 3 | 3 (0.33) | 2 (0.22) | 5 (0.28) | 0.6858 |
| Diarrhea | 2 (0.22) | 2 (0.22) | 4 (0.22) | 1.0000 |
| Grade 1 | 2 (0.22) | 2 (0.22) | 4 (0.22) | 1.0000 |
| Vomiting | 2 (0.22) | 1 (0.11) | 3 (0.17) | 0.6231 |
| Grade 1 | 1 (0.11) | 1 (0.11) | 2 (0.11) | 1.0000 |
| Grade 2 | 1 (0.11) | 0 | 1 (0.06) | 0.4978 |
| Nausea | 5 (0.56) | 2 (0.22) | 7 (0.39) | 0.2861 |
| Grade 1 | 4 (0.45) | 2 (0.22) | 6 (0.33) | 0.4504 |
| Grade 2 | 1 (0.11) | 0 | 1 (0.06) | 0.4978 |
| Muscle pain (non-inoculation site) | 30 (3.35) | 25 (2.77) | 55 (3.06) | 0.4961 |
| Grade 1 | 19 (2.12) | 16 (1.77) | 35 (1.94) | 0.6128 |
| Grade 2 | 10 (1.12) | 8 (0.88) | 18 (1.00) | 0.6446 |
| Grade 3 | 1 (0.11) | 1 (0.11) | 2 (0.11) | 1.0000 |
| Joint pain | 2 (0.22) | 2 (0.22) | 4 (0.22) | 1.0000 |

|  |  | **NVSI-06-08**<br>**(N=896)** | **BBIBP-CorV**<br>**(N=904)** | **Total**<br>**(N=1800)** | **P value\*** |
| --- | --- | --- | --- | --- | --- |
|  | Grade 1 | 1 (0.11) | 2 (0.22) | 3 (0.17) | 1.0000 |
|  | Grade 2 | 1 (0.11) | 0 | 1 (0.06) | 0.4978 |
|  | Headache | 52 (5.80) | 38 (4.20) | 90 (5.00) | 0.1304 |
|  | Grade 1 | 30 (3.35) | 18 (1.99) | 48 (2.67) | 0.0801 |
|  | Grade 2 | 23 (2.57) | 20 (2.21) | 43 (2.39) | 0.6463 |
|  | Cough | 20 (2.23) | 13 (1.44) | 33 (1.83) | 0.2233 |
|  | Grade 1 | 14 (1.56) | 9 (1.00) | 23 (1.28) | 0.3024 |
|  | Grade 2 | 6 (0.67) | 5 (0.55) | 11 (0.61) | 0.7728 |
|  | Breathing trouble | 0 | 1 (0.11) | 1 (0.06) | 1.0000 |
|  | Grade 1 | 0 | 1 (0.11) | 1 (0.06) | 1.0000 |
|  | Fatigue | 27 (3.01) | 22 (2.43) | 49 (2.72) | 0.4721 |
|  | Grade 1 | 19 (2.12) | 17 (1.88) | 36 (2.00) | 0.7393 |
|  | Grade 2 | 9 (1.00) | 5 (0.55) | 14 (0.78) | 0.2979 |
|  | Dizziness | 1 (0.11) | 2 (0.22) | 3 (0.17) | 1.0000 |
|  | Grade 1 | 1 (0.11) | 1 (0.11) | 2 (0.11) | 1.0000 |
|  | Grade 2 | 0 | 1 (0.11) | 1 (0.06) | 1.0000 |
|  | **Unsolicited adverse reactions within 0-30 days** | 48 (5.36) | 48 (5.31) | 96 (5.33) |  |
|  | Grade 1 | 40 (4.46) | 32 (3.54) | 72 (4.00) |  |
|  | Grade 2 | 13 (1.45) | 22 (2.43) | 35 (1.94) |  |
|  | Grade 3 | 2 (0.22) | 1 (0.11) | 3 (0.17) |  |

\* P-value was calculated using Fisher's exact test. Data are presented as n (%).

**Extended Data Table 3.** Live-virus neutralizing antibody response results (PPS)

|  | 15 days after boosting |  |  | 30 days after boosting |  |  |
| --- | --- | --- | --- | --- | --- | --- |
|  | NVSI-06-08 | BBIBP-CorV | P value | NVSI-06-08 | BBIBP-CorV | P value |
| 4-6 months |  |  |  |  |  |  |
| N(missing) | 285 (0) | 289 (0) |  | 285 (0) | 289 (0) |  |
| Pre-booster antibody GMT<sup>[1]</sup> (95%CI) | 78.35 (67.10-91.48) | 67.28 (57.41-78.84) | 0.1771 | 78.35 (67.10-91.48) | 67.28 (57.41-78.84) | 0.1771 |
| Post-booster antibody GMT (95%CI) | 3141.92 (2731.70-3613.74) | 196.89 (176.01-220.24) | <0.0001 | 3730.18 (3216.15-4326.36) | 294.96 (266.23-326.80) | <0.0001 |
| Ratio of GMT between two groups(95%CI)<sup>[2]</sup> | 15.96 (13.34-19.08) |  |  | 12.65 (10.56-15.14) |  |  |
| Rate of 4-fold rise<sup>[3]</sup>, n (%) | 267 (93.68) | 66 (22.84) | <0.0001 | 276 (96.84) | 112 (38.75) | <0.0001 |
| 95%CI (%) | 90.20-96.21 | 18.13-28.12 |  | 94.09-98.55 | 33.11-44.64 |  |
| Rate difference between two groups (% , 95%CI<sup>[4]</sup>) | 70.85 (64.82-76.03) |  |  | 58.09 (51.92-63.84) |  |  |
| Post-booster antibody GMT fold rise (95%CI) | 40.10 (34.61-46.47) | 2.93 (2.54-3.37) | <0.0001 | 47.61 (41.17-55.06) | 4.38 (3.81-5.05) | <0.0001 |
| 7-9 months |  |  |  |  |  |  |
| N(missing) | 270 (0) | 270 (0) |  | 270 (0) | 270 (0) |  |
| Pre-booster antibody GMT<sup>[1]</sup> (95%CI) | 51.98 (44.89-60.19) | 48.80 (42.03-56.66) | 0.5526 | 51.98 (44.89-60.19) | 48.80 (42.03-56.66) | 0.5526 |
| Post-booster antibody GMT (95%CI) | 4908.34 (4259.82-5655.59) | 504.76 (443.94-573.92) | <0.0001 | 7719.35 (6828.68-8726.20) | 1093.26 (958.59-1246.84) |  |
| Ratio of GMT between two groups(95%CI)<sup>[2]</sup> | 9.72 (8.04-11.77) |  |  | 7.06 (5.90-8.45) |  |  |
| Rate of 4-fold rise<sup>[3]</sup>, n (%) | 265 (98.15) | 203 (75.19) | <0.0001 | 268 (99.26) | 251 (92.96) | 0.0002 |
| 95%CI (%) | 95.73-99.40 | 69.59-80.22 |  | 97.35-99.91 | 89.23-95.71 |  |
| Rate difference between two groups (% , 95%CI<sup>[4]</sup>) | 22.96 (17.80-28.63) |  |  | 6.30 (3.08-9.51) |  |  |
| Post-booster antibody GMT fold rise (95%CI) | 94.42 (79.36-112.34) | 10.34 (8.78-12.19) | <0.0001 | 148.50 (126.60-174.19) | 22.40 (19.19-26.15) | <0.0001 |
| >9 months |  |  |  |  |  |  |
|  | NVSI-06-08 | BBIBP-CorV | P value | NVSI-06-08 | BBIBP-CorV | P value |
| N(missing) | 286 (0) | 278 (0) |  | 286 (0) | 278 (0) |  |
| Pre-booster antibody GMT<sup>[1]</sup> (95%CI) | 16.96 (14.56-19.74) | 17.47 (14.94-20.44) | 0.7870 | 16.96 (14.56-19.74) | 17.47 (14.94-20.44) | 0.7870 |
| Post-booster antibody GMT (95%CI) | 4184.82 (3651.45-4796.10) | 374.63 (339.20-413.76) | <0.0001 | 7479.16 (6540.06-8553.10) | 808.29 (714.16-914.82) | <0.0001 |
| Ratio of GMT between two groups(95%CI)<sup>[2]</sup> | 11.17 (9.44-13.22) |  |  | 9.25 (7.71-11.10) |  |  |
| Rate of 4-fold rise<sup>[3]</sup>, n (%) | 285 (99.65) | 262 (94.24) | <0.0001 | 286 (100.00) | 273 (98.20) | 0.0286 |
| 95%CI (%) | 98.07-99.99 | 90.82-96.67 |  | 98.72-100.00 | 95.85-99.41 |  |
| Rate difference between two groups (% , 95%CI<sup>[4]</sup>) | 5.41 (2.96-8.84) |  |  | 1.80 (0.46-4.14) |  |  |
| Post-booster antibody GMT fold rise (95%CI) | 246.81 (207.02-294.26) | 21.44(18.56-24.77) | <0.0001 | 441.11 (373.91-520.38) | 46.26 (39.76-53.83) | <0.0001 |

Notes: [1] GMT represent geometric mean titer.
[2] The ratio of GMT between two groups was calculated by “NVSI-06-08/ BBIBP-CorV”, and the non-inferiority threshold of ratio between groups on day 15 post-booster was set to 0.67.
[3] Rate of 4-fold rise was defined as percentage of participants with a $\geq 4$ -fold rise from baseline in neutralizing antibody titer.
[4] Rate difference=(NVSI-06-08)-(BBIBP-CorV). Rate difference and 95%CI were estimated by Miettinen-Nurminen method without considering stratification factors.

**Extended Data Table 4.** RBD-specific IgG response results (PPS)

|  | 15 days after boosting |  |  | 30 days after boosting |  |  |
| --- | --- | --- | --- | --- | --- | --- |
|  | NVSI-06-08 | BBIBP-CorV | P value | NVSI-06-08 | BBIBP-CorV | P value |
| **4-6 months** |  |  |  |  |  |  |
| N(missing) | 285 (0) | 289 (0) |  | 285 (0) | 289 (0) |  |
| Pre-booster antibody GMC<sup>[1]</sup> (95%CI) | 113.71 (96.45-134.05) | 95.53 (81.53-111.94) | 0.1341 | 113.71 (96.45-134.05) | 95.53 (81.53-111.94) | 0.1341 |
| Post-booster antibody GMC (95%CI) | 7119.99 (6235.45-8130.01) | 254.00 (228.22-282.69) | <0.0001 | 5030.67 (4416.40-5730.38) | 213.91 (191.73-238.66) | <0.0001 |
| Ratio of GMC between two groups (95%CI)<sup>[2]</sup> | 28.03(23.65-33.23) |  |  | 23.52 (19.85-27.87) |  |  |
| Rate of 4-fold rise<sup>[3]</sup>, n (%) | 274 (96.14) | 60 (20.76) | <0.0001 | 272 (95.44) | 50 (17.30) | <0.0001 |
| 95%CI (%) | 93.20-98.06 |  |  | 92.33-97.55 | 13.12-22.16 |  |
| Rate difference between two groups (% , 95%CI<sup>[4]</sup>) | 75.38 (69.76-80.13) |  |  | 78.14 (72.67-82.66) |  |  |
| Post-booster antibody GMC fold rise (95%CI) | 62.62 (53.00-73.98) | 2.66(2.30-3.08) | <0.0001 | 44.24 (37.82-51.75) | 2.24 (1.94-2.58) | <0.0001 |
| **7-9 months** |  |  |  |  |  |  |
| N(missing) | 270 (0) | 270 (0) |  | 270 (0) | 270 (0) |  |
| Pre-booster antibody GMC<sup>[1]</sup> (95%CI) | 133.93 (111.27-161.20) | 118.77 (97.65-144.47) | 0.3810 | 133.93 (111.27-161.20) | 118.77 (97.65-144.47) | 0.3810 |
| Post-booster antibody GMC (95%CI) | 6583.20 (5734.58-7557.39) | 303.96 (270.91-341.04) | <0.0001 | 4947.49 (4395.20-5569.19) | 290.61 (258.00-327.35) | <0.0001 |
| Ratio of GMC between two groups (95%CI)<sup>[2]</sup> | 21.66 (18.10-25.91) |  |  | 17.02 (14.40-20.13) |  |  |
| Rate of 4-fold rise<sup>[3]</sup>, n (%) | 250 (92.59) | 62 (22.96) | <0.0001 | 249 (92.22) | 62 (22.96) | <0.0001 |
| 95%CI (%) | 88.79-95.42 | 18.08-28.45 |  | 88.36-95.12 | 18.08-28.45 |  |
| Rate difference between two groups (% , 95%CI<sup>[4]</sup>) | 69.6 3 (63.27-75.10) |  |  | 69.26 (62.87-74.76) |  |  |
| Post-booster antibody GMC fold rise (95%CI) | 49.15 (40.43-59.76) | 2.56 (2.13-3.07) | <.0001 | 36.94 (30.65-44.52) | 2.45 (2.04-2.93) | <.0001 |
| **>9 months** |  |  |  |  |  |  |
| N(missing) | 286(0) | 278(0) |  | 286(0) | 278(0) |  |
|  | NVSI-06-08 | BBIBP-CorV | P value | NVSI-06-08 | BBIBP-CorV | P value |
| Pre-booster antibody GMC<sup>[1]</sup> (95%CI) | 120.45 (96.96-149.63) | 116.05 (91.23-147.62) | 0.8210 | 120.45 (96.96-149.63) | 116.05 (91.23-147.62) | 0.8210 |
| Post-booster antibody GMC (95%CI) | 7440.80 (6509.03-8505.96) | 414.87 (362.17-475.25) | <0.0001 | 4960.67 (4379.76-5618.63) | 267.88 (233.41-307.44) | <0.0001 |
| Ratio of GMC between two groups (95%CI)<sup>[2]</sup> | 17.94 (14.83-21.69) |  |  | 18.52 (15.39-22.29) |  |  |
| Rate of 4-fold rise<sup>[3]</sup>, n (%) | 259 (90.56) | 88 (31.65) | <0.0001 | 252 (88.11) | 61 (21.94) | <0.0001 |
| 95%CI (%) | 86.56-93.69 | 26.23-37.48 |  | 83.79-91.62 | 17.22-27.27 |  |
| Rate difference between two groups (% , 95%CI<sup>[4]</sup>) | 58.90 (52.14-65.00) |  |  | 66.17 (59.59-71.88) |  |  |
| Post-booster antibody GMC fold rise (95%CI) | 61.77 (49.74-76.72) | 3.58 (2.93-4.36) | <0.0001 | 41.18 (33.25-51.01) | 2.31 (1.89-2.82) | <0.0001 |

Notes: [1] GMC represents geometric mean concentration, and the unit of GMC is BAU/ml.
[2] The ratio of GMC between two groups was calculated by “NVSI-06-08/ BBIBP-CorV”.
[3] Rate of 4-fold rise was defined as percentage of participants with a $\geq 4$ -fold rise from baseline in IgG concentration.
[4] Rate difference=(NVSI-06-08)-(BBIBP-CorV). Rate difference and 95%CI were estimated by Miettinen-Nurminen method without considering stratification factors.

**Extended Data Table 5.**
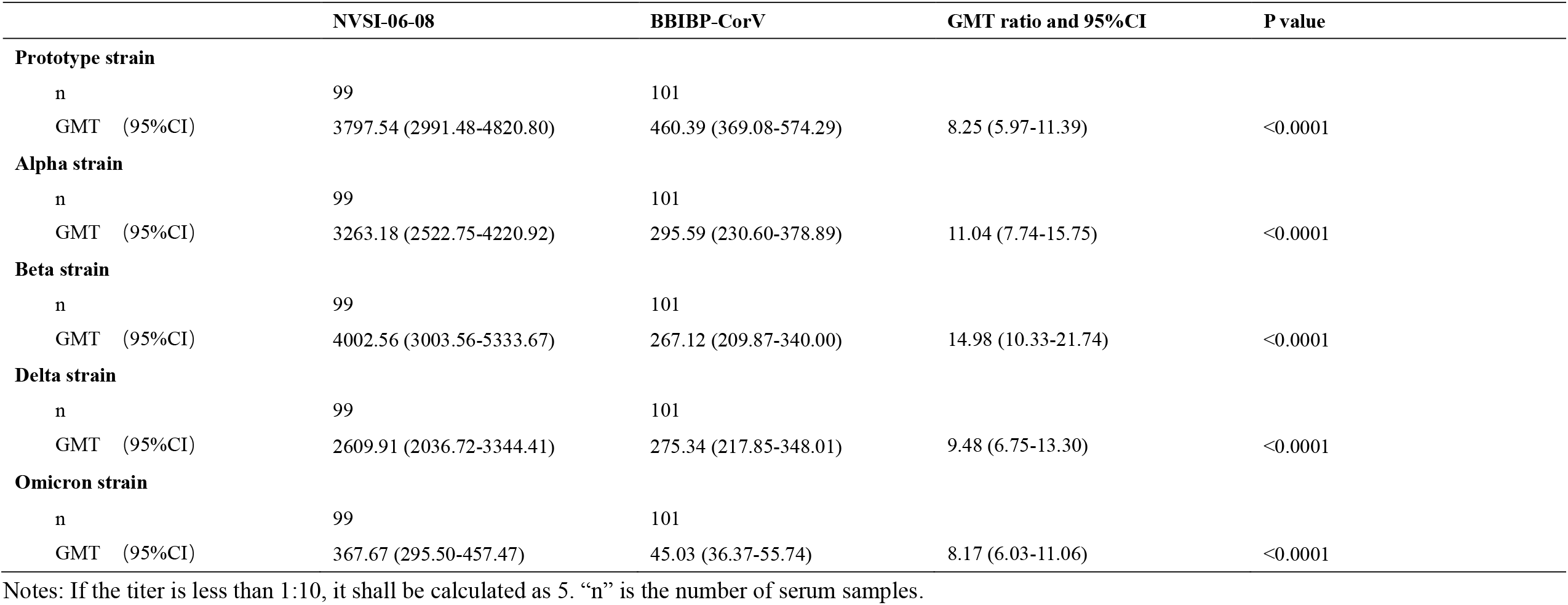
Live-virus neutralizing antibody responses against main SARS-CoV-2 VOC variants.

|  | NVSI-06-08 | BBIBP-CorV | GMT ratio and 95%CI | P value |
| --- | --- | --- | --- | --- |
| **Prototype strain** |  |  |  |  |
| n | 99 | 101 |  |  |
| GMT (95%CI) | 3797.54 (2991.48-4820.80) | 460.39 (369.08-574.29) | 8.25 (5.97-11.39) | <0.0001 |
| **Alpha strain** |  |  |  |  |
| n | 99 | 101 |  |  |
| GMT (95%CI) | 3263.18 (2522.75-4220.92) | 295.59 (230.60-378.89) | 11.04 (7.74-15.75) | <0.0001 |
| **Beta strain** |  |  |  |  |
| n | 99 | 101 |  |  |
| GMT (95%CI) | 4002.56 (3003.56-5333.67) | 267.12 (209.87-340.00) | 14.98 (10.33-21.74) | <0.0001 |
| **Delta strain** |  |  |  |  |
| n | 99 | 101 |  |  |
| GMT (95%CI) | 2609.91 (2036.72-3344.41) | 275.34 (217.85-348.01) | 9.48 (6.75-13.30) | <0.0001 |
| **Omicron strain** |  |  |  |  |
| n | 99 | 101 |  |  |
| GMT (95%CI) | 367.67 (295.50-457.47) | 45.03 (36.37-55.74) | 8.17 (6.03-11.06) | <0.0001 |

Notes: If the titer is less than 1:10, it shall be calculated as 5. “n” is the number of serum samples.

